# Genome-wide association study of pediatric bacteremia and sepsis

**DOI:** 10.1101/2025.06.12.25329506

**Authors:** Dylan Lawless, Flavia Aurelia Hodel, Christian W. Thorball, Zhi Ming Xu, Alessandro Borghesi, Eric Giannoni, Johannes Trück, Martin Stocker, Klara M Posfay-Barbe, Ulrich Heininger, Sara Bernhard-Stirnemann, Anita Niederer-Loher, Christian R. Kahlert, Giancarlo Natalucci, Christa Relly, Christoph Berger, Thomas Riedel, Christoph Aebi, Philipp Agyeman, Jacques Fellay, Luregn J. Schlapbach, the Swiss Pediatric Sepsis Study

## Abstract

**Background:** Sepsis is defined as a dysregulated host response to infection leading to organ dysfunction. It represents a major global health concern, particularly in childhood. The precise underlying pathophysiological mechanisms remain poorly understood.

**Methods:** Using samples and clinical data from 650 children enrolled in the Swiss Pediatric Sepsis Study, a national multicenter cohort study for culture-proven bacterial sepsis, and from 1395 controls, we explored the human genetic determinants of inter-individual variability in sepsis-related outcomes. We used a case-control study design to search for associations between genome-wide polymorphisms and susceptibility to sepsis, and case-only analysis for specific characteristics of the disease.

**Findings:** We identified one locus significantly associated with sepsis susceptibility, encompassing the *CTNNAL1* and *ELP1* genes.

**Interpretation:** Our results suggest a genetic modulator of sepsis susceptibility in children.

**Funding:** This study was funded by grants from the Swiss National Science Foundation (320030_201060 and 342730 153158/1), the Swiss Society of Intensive Care, the Bangerter Foundation, the Vinetum and Borer Foundation, and the Foundation for the Health of Children and Adolescents.

**Research in context:** *Evidence before this study:* Blood culture-confirmed bacterial sepsis in children remains an important cause of morbidity and mortality. Several prospective studies from the Swiss Pediatric Sepsis Study (SPSS) have defined its incidence, clinical features, and outcomes in Switzerland. A 2017 nationwide cohort study reported an incidence of 25.1 per 100,000 children annually and a 30-day mortality rate of 7%, with risk of death closely associated with the presence and number of organ dysfunctions. A neonatal-focused analysis identified three distinct clinical subgroups (early-onset, hospital-acquired, and community-acquired late-onset sepsis), each with specific pathogens, risk profiles, and outcomes. Hospital-acquired infections were most frequent and carried the highest case fatality. In a separate cohort of previously healthy children with community-acquired sepsis, rare variants in primary immunodeficiency genes were identified in 20% of cases, although most were of uncertain significance. Another SPSS study assessed organ dysfunction scores in paediatric sepsis and found similar performance in predicting 30-day mortality. Cardiovascular, respiratory, and neurologic dysfunctions were most strongly associated with death, suggesting that simplified assessment may be feasible.

*Added value of this study:* This is the first genome-wide association study of paediatric bacterial sepsis based on a national cohort with blood culture-confirmed diagnoses. The cohort included 650 sepsis cases and 1395 population-based controls. A genome-wide significant susceptibility locus was identified on chromosome 9 (CTNNAL1/ELP1). Some biological relevance of this locus has previously been demonstrated independently. The study expands previous SPSS research by integrating epidemiological data with genome-wide genetics and by applying stringent ancestry correction and imputation within the clinical phenotype. The findings offer new avenues for investigating genetic contributions to paediatric sepsis susceptibility.

*Implications of all the available evidence:* Combined with earlier SPSS studies, these findings support a model in which both genetic and clinical factors contribute to sepsis susceptibility and outcome in children. The identification of a susceptibility locus near CTNNAL1 and ELP1 provides a new basis for mechanistic studies and implicates transcriptional regulation and NF-κB signalling pathways. Use of a microbiologically confirmed phenotype enhances interpretability and relevance for translational research. These results provide a foundation for future work on host-pathogen interactions, genetic risk stratification, and targeted interventions in paediatric sepsis.

## Introduction

Sepsis represents a leading cause of death and disability in adults and children [1], [2], [3]. Globally, pediatric sepsis results in over one million annual deaths [4], [5]. Even in countries with low childhood mortality, sepsis contributes to 10 to 20% of childhood deaths, and to one out of four deaths in pediatric intensive care units (PICU) [6], [7]. Sepsis is the result of a dysregulated host response to infection leading to organ dysfunction, yet the biomolecular mechanisms and host-pathogen interactions involved remain poorly understood [8], [9], [10]. A key feature of sepsis is that it usually develops after an insidious onset of non-specific signs common to many mild infections before progressing to organ dysfunction. In clinical practice, recognizing this transition from an infected child who may solely require antibiotics to a septic child progressing to multiorgan failure (MOF) and death, is extremely challenging.

The Swiss Pediatric Sepsis Study (SPSS) is a national observational multicenter cohort study investigating blood culture-proven bacterial sepsis in children in Switzerland. Clinical and laboratory data have been have been previously reported [2], [11], [12]. We have also previously reported population-based incidence estimates and findings on the epidemiology of sepsis [1]. Based on our data, 4.4% (95%-CI 3.5-5.4) of all childhood deaths in Switzerland were associated with blood culture-proven bacterial sepsis, ranking 5^th^ as the cause of all childhood deaths. The age-standardized incidence of sepsis was 25.1 per 100’000 children per year with an average 30-day in-hospital mortality of 6.9% (95%-CI 5.6-8.6), with highest rates observed in children under five years of age.

The incidence of sepsis is highest during early life [13], a vulnerable period of adaptive immunity development [14], [15], [16]. The mechanisms underlying susceptibility and resistance to severe infectious diseases in otherwise healthy patients remains largely unknown, however epidemiological studies suggest a strong genetic contribution [17], [18]. Previous studies have used mostly candidate gene and linkage analyses [19]. Genome-wide association studies (GWAS), which allow a comprehensive interrogation of potential associations throughout the genome, have identified common genetic variants that are associated with many infectious diseases [20]. Understanding why a minority of children become very ill during the course of an infection remains one of the unsolved riddles in host genetic research, and may harbor clues for future targeted interventions. We here use a GWAS approach to elucidate host genetic determinants of susceptibility to pediatric sepsis and disease characteristics.

## Methods

### Sample collection

The SPSS ran prospectively from September 1, 2011, to December 31, 2015. During the recruitment period, participating centers accounted for 77.6% of all hospital admissions and 97.8% of all pediatric intensive care unit admissions with an ICD-10 code for pathogen-specific sepsis in children below the age of 17 years in Switzerland. A total of 1269 blood culture-proven sepsis episodes were recorded in 1164 children. Detailed clinical and laboratory data were collected to assess predictors including pediatric organ dysfunction scores [2], [3], [4].

### Genotype quality control and imputation

Genomic DNA was extracted from whole blood of 704 participants. From these, 670 were patients with clinical features and genomic data suitable for our analysis. Samples were genotyped using Illumina OmniExpressExome-8 v1.4 genotyping array and genotypes were called using Illumina GenomeStudio. An in-house control cohort of 1395 Swiss population samples was included. Study participants were excluded based on a missing genotype call rate of 10%. Subject independence was assessed using KING; nine samples were removed due to a high degree of kinship or duplication (pairwise identify-by-state (IBS) estimated kinship coefficient > 0.18) [21]. From the distribution of genetic distance, no exclusion of outliers was necessary. One sample was removed based on retraction of genetic consent.

Variants were removed for minor allele frequencies < 0.05, missingness > 0.1, and additionally for controls, Hardy-Weinberg Equilibrium (HWE) P < 1E-6. Reported and estimated sex was examined for discrepancy. We compared the genetic ancestry in cases to self-reported ethnicity to check for mislabeling. After quality control (QC), 650 sepsis case samples and 1395 controls remained. Genotyping data was phased (SHAPEIT2) and imputed (IMPUTE2) using the 1000 Genomes Project phase 3 reference panel. The reference genome build and LD population used was hg19/1000G Nov2014 EUR. Imputation quality was assessed and SNPs with an information score of < 0.8 or minor allele frequency < 0.05 were removed (**Figure S2**).

### Association testing

GCTA was used to calculate the genetic relationship matrix (GRM) and to perform principal component analysis (PCA) to quantify population structure [22]. For case-control analysis of sepsis susceptibility, population structure/stratification was alleviated by sub-setting the dataset to remove outlying clusters as evident from PCA (**Figure S3**). This removed 140 cases and 401 controls, leaving a case-control subset of 510 cases and 994 controls of European ancestry. Datasets were merged using PLINK v1.9. SNP positions and identifiers were updated according to dbNSFP4.0a (hg19) [23]. QC was repeated after merging cases and controls for combined cohort-specific frequencies and GCTA mixed linear model-based association (MLMA) with leaving-one-chromosome-out (LOCO) was used for analysis. SNP QC filters and association model summaries are listed in **Tables S1 and S2**, respectively. GCTA MLMA-LOCO was used for within-cohort (case only) analyses of several phenotypes as listed in **Table 1 and S2** [24]. Population structure was controlled by GRM eigenvectors and analysis covariates consisted of sex, age, and study site.

**Table 1:** Demographic and clinical characteristics of 650 Swiss Pediatric Sepsis Study GWAS participants. Categorical variables are presented as frequencies (%) and continuous variables as median (IQR). Column percentages are presented based on available data for each variable. ^a^Missing in 19, ^b^Missing in 1, ^c^Missing in 1, ^d^Missing in 3, ^e^Missing in 9, ^f^In 3 cases laboratory and physiological parameters needed to define organ dysfunctions according to the 2005 consensus definition were missing, but according to the initial investigator-based classification these patients did not suffer from any organ dysfunction. CVAD, central venous access device; PICU, paediatric intensive care unit; NICU, neonatal intensive care unit.

### Testing for overlap with an independent mouse model study

We identified *ELP1* and *CTNNAL1* as candidate genes for susceptibility to paediatric sepsis. These genes (*Elp1* and *Ctnnal1* in mice) were independently reported by Vered et al. [26] as part of the Collaborative Cross (CC), a systems genetics resource for host-pathogen interaction studies using a multi-parental mouse genetic reference population, as reviewed by Noll et al. [25]. Vered et al. [26] challenged 73 pre-CC mouse lines with *Klebsiella pneumoniae* and identified 2 and 6 significantly associated intronic SNPs in Elp1 and Ctnnal1, respectively. To test whether the overlap of these genes across the two studies exceeded that expected by chance, we applied a weighted Z-test and Fisher’s combined probability test using the reported study sizes (n=1504 for SPSS and n=328 for CC).

### Ethics statement

The study was approved by the respective ethics committees of all participating centers (Cantonal Ethics Committee Bern, approval number KEK-029/11) and the study was conducted in accordance with the Declaration of Helsinki. Participants or their legally acceptable representatives provided their written consent to participate in the study after having been informed about the nature and purpose of the study, participation/termination conditions, and risks and benefits of treatment.

### Role of funders

The funders were not involved in study design, data collection and analysis, interpretation, and writing of the manuscript.

## Results

The demographic and clinical characteristics of the 650 study participants for whom within-cohort analysis was performed are shown in **Table 1** and **Figure S1**. Within-cohort (case only) association testing was performed on all 650 case samples. We found no association with specific sepsis characteristics, including site or type of infection, cardiovascular failure, comorbidities, pathogens identified, case fatality, PICU admission, hospital-acquired sepsis, length of PICU stay, time to death from sepsis onset, number of organ dysfunctions, length of stay after sepsis onset, invasive ventilation, and age groups (**Table S2**). The sub-group summaries are listed with demographic and clinical characteristics in **Table 1** and **Figure S1**.

A subset of 510 cases and 994 Swiss population-matched controls were included in a case-control GWAS for sepsis susceptibiliy as shown in **Figure 1**. This analysis highlighted one significantly associated region of chromosome 9 with 13 genome-wide significant SNPs and rs28361152 as the top-associated variant (**Table 2** and **Figure 2A**). Conditional analysis with GCTA-COJO showed that all SNPs reflect the same association signal. All 13 SNPs were intronic and did not initially appear to affect protein structure, but deeper analyses revealed their functional effects as expression Quantitative Trait Loci (eQTL). The QQ-plot illustrated in **Figure 2B** indicates that genomic inflation was controlled.

**Figure 1:**
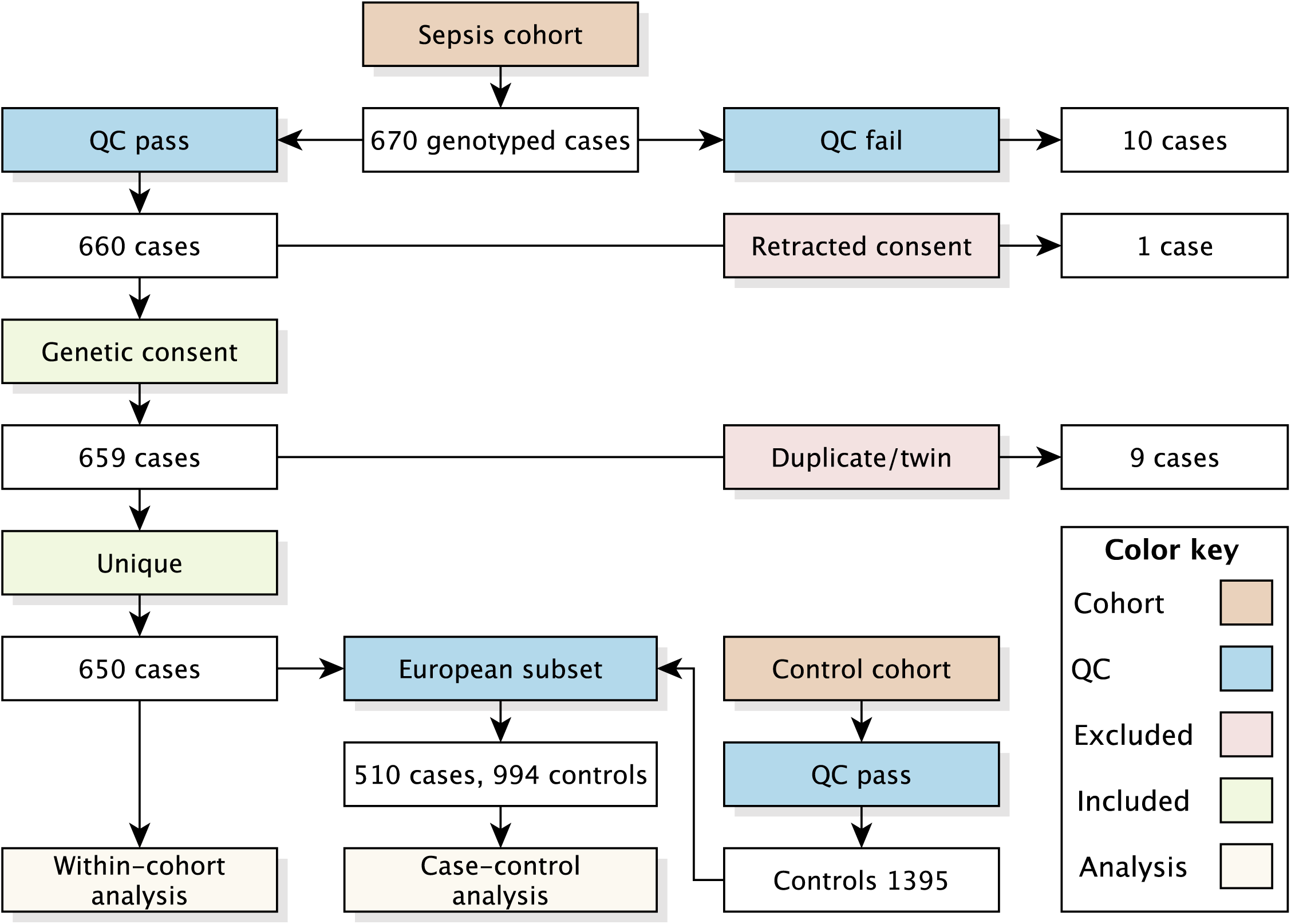
Cohort analysis summary. Swiss Pediatric Sepsis cohort episodes, genotyping, and quality control. Within-cohort analysis are listed in Table S2. Case-control analysis subset to suitably match cohorts.

**Figure 2:**
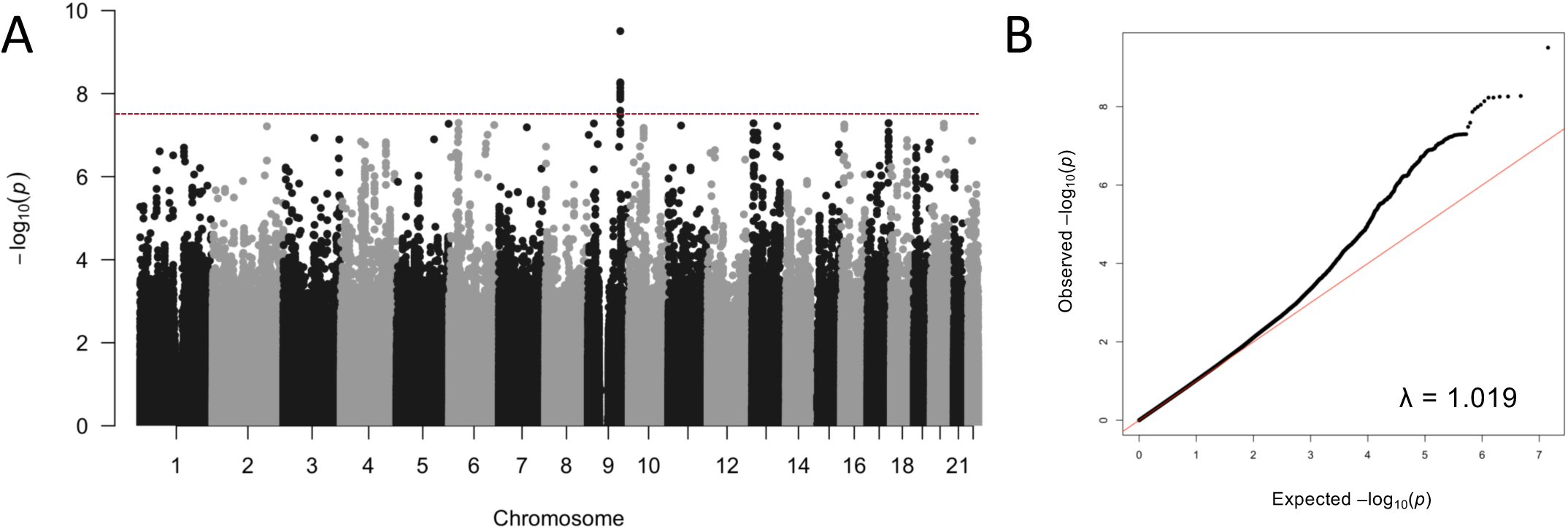
GWAS Manhattan plot and QQ plot. (A) Manhattan plot of association shows significant association signals in chromosome 9 (significant threshold P-value = 5e^−08^). (B) QQ plot demonstrates that the observed distribution of P-values corresponds to the expected distribution under the null hypothesis, indicating that potential confounders are well controlled, λ=1.019.

**Table 2.** Significant associations identified for case and control susceptibility. Lead SNP rs28361152 is a non-coding variant that falls within *CTNNAL1*. The remaining associated SNPs in LD (r^2^ ≥ 0.8) span *ELP1* and *FAM206A*.

The lead SNP, associated variants and gene positions in this region are shown in a LocusZoom plot in **Figure 3**. The strength and extent of the association signal relative to genomic position, local LD and recombination patterns indicate five genes of potential interest: *ELP1* (elongator acetyltransferase complex subunit 1), *FAM206A* (family with sequence similarity 206, member A), *CTNNAL1* (catenin alpha-like 1 gene), *TMEM245* (Transmembrane protein 245), and *FRRS1L* (DOMON domain-containing protein FRRS1L). Using GTEx v.8 data (**Figure S4**), we observed that the top associated variant is a significant cis-eQTL for all five genes in the LD block (**Figure 3**).

**Figure 3:**
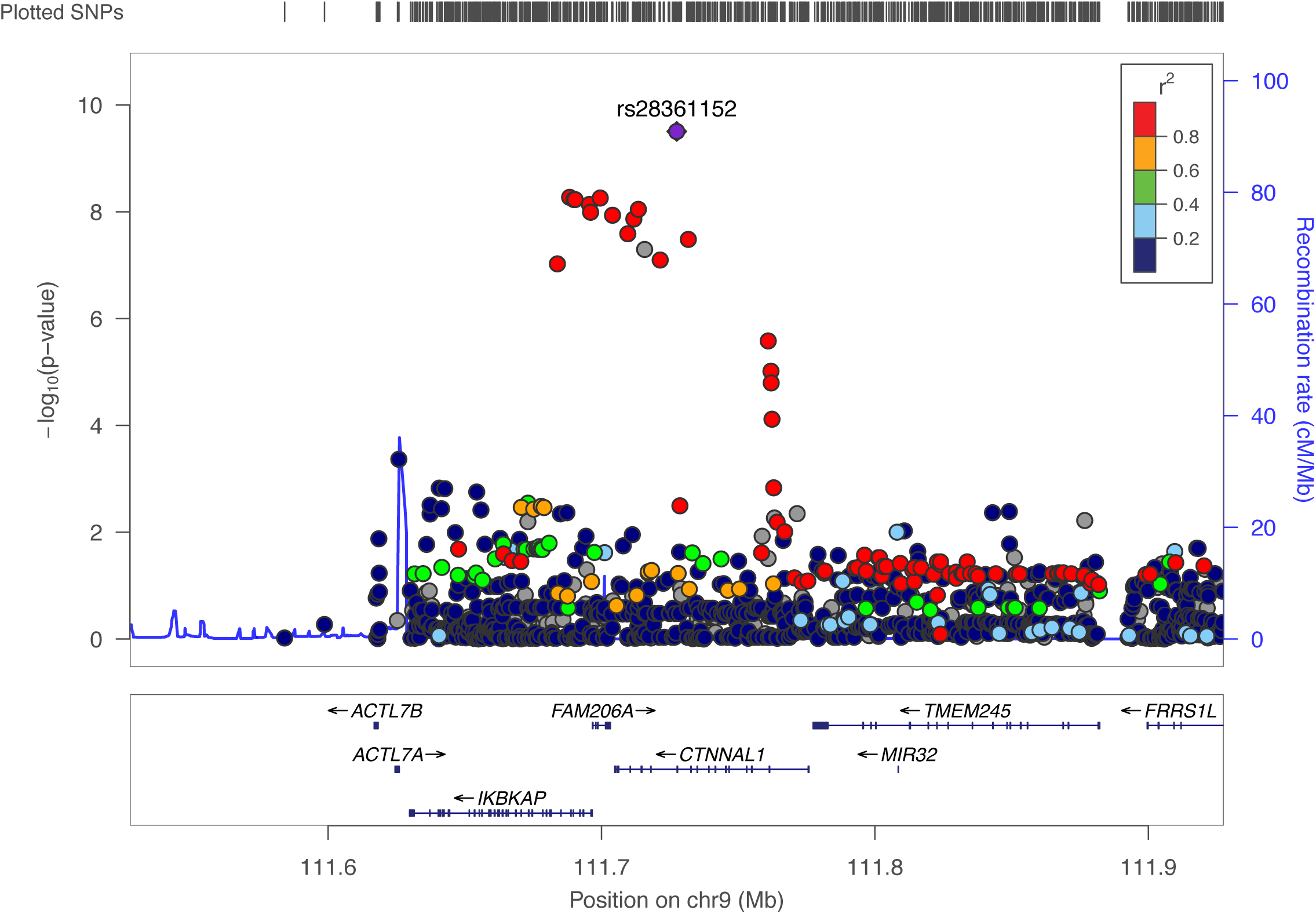
A LocusZoom plot for SNPs in the region flanking 200kb on either side of the selected SNP rs28361152 on chromosome 9. P-values in –log10 scale are shown on the left vertical axis, the recombination rates are on the right vertical axis, and the chromosomal positions are on the horizontal axis. LD is shown by r^2^ and represented by color. The lead SNP is in strong LD with nearest gene *CTNNAL1*, *FAM206A,* and *ELP1.* The LD block extends to *TMEM245* and *FRRS1L,* but no associated with disease is evident within the SPSS cohort.

The details of all coding and non-coding variants in LD are provided in **Table S3** and **Table S4**, respectively. **Table S5** shows all significant eQTLs for the 13 SNPs associated with susceptibility to sepsis in our study. The strongest association between rs28361152 and mRNA expression differences was found for *FRRS1L*, approximately 165kb downstream of rs28361152 with increased expression in presence of the minor allele. rs28361152 was also found to act as a splicing Quantitative Trait Locus (sQTL) in *ELP1* for intron ID 108906485:108908305:clu_63656 (GRCh38) affecting exon 14 ENSE00000983463 and intron 14-15 (**Table S6**). All genes in the region are highly expressed in most tissues, with the exception of *FRRS1L*, which has low expression globally (**Figure S4**).

*ELP1* and *CTNNAL1* (and murine *Elp1* and *Ctnnal1*) were identified as candidate causal genes in both our paediatric sepsis susceptibility scan and in a previously reported independent scan of 73 mouse lines challenged with *K. pneumoniae* by Vered et al. [26], in the CC study [25]. *K. pneumoniae* causes severe pneumonia and sepsis in immunocompromised individuals. We tested the probability of identifying the same two genes in the two independent scans and found it highly unlikely to be due to chance: P-value=1.28E-13 using weighted *Z*-test (study sizes 1504 and 328 for SPSS and CC, respectively), and 1.53E-17 using Fisher’s combined probability test. Infection with *Klebsiella* spp. was reported in 28 of our 510 sepsis cases. The direction and strength of the association with rs28361152 was comparable between sepsis due to *Klebsiella* spp. (OR 0.68, 95% CI 0.3-1.56) and sepsis due to other pathogens (OR 0.46, 95% CI 0.37-0.58).

## Discussion

As part of the Swiss Pediatric Sepsis Study, we explored culture-proven bacteremia and sepsis-related outcomes for 650 cases and performed case-control analysis for 510 cases and 994 controls of European ancestry. Our search for human genetic determinants of susceptibility to pediatric sepsis identified a locus of interest on chromosome 9 with 13 genome-wide significant SNPs. The top associated variant, rs28361152, is an eQTL for five genes in the immediate vicinity - *ELP1, FAM206A*, *CTNNAL1*, TMEM245, and *FRRS1L*. It is also an sQTL for *ELP1*. Of note, *elp1* and *ctnnal1* have been previously reported as top candidate genes in a murine model of sepsis [26].

*CTNNAL1* encodes alpha-catulin, a regulatory protein that is associated with inflammatory and immune pathways through modulation of the NF-kB pathway activity [27]. Murine Ctnnal1 is also important to cell adhesion, in response to bacterial triggers, to eliminate infection [28], [29]. *ELP1* encodes the elongator complex protein 1, a component of the RNA polymerase II elongator complex, which is involved in transcriptional elongation. This complex catalyzes the formation of carboxymethyluridine in the wobble base at position 34 in tRNAs [30]. Loss of mouse *Elp1* is embryonic lethal and can be rescued by human *ELP1* [31].

The Swiss Pediatric Sepsis Study addressed the pressing need for comprehensive, longitudinal, population-based cohorts in paediatric sepsis - a condition that remained a leading cause of death and disability globally [5]. The relatively small sample size limited the power of some analyses. In particular, the case-only analyses were hindered by insufficient sample numbers, and the case-control analysis was constrained by the limited availability of well-matched controls for non-European genetic backgrounds.

Differences in allele frequencies further highlighted the challenges in this research area. The in-house Swiss population control cohort revealed a minor allele frequency of approximately 0.3 for the lead variant, consistent with data from gnomAD and FinnGen. In contrast, African and South Asian populations exhibited much lower frequencies, 0.05 and 0.0, respectively, suggesting that sepsis susceptibility might have varied significantly with ancestry. Increasing sample sizes and increasing the controls in future studies could enhance the ability to detect additional genetic associations more effectively.

Although GWAS provided valuable statistical associations, they fall short of pinpointing causal variants, especially in non-coding regions where genomic complexity is a major hurdle. Replication studies with independent cohorts, along with functional assays and fine mapping, are preferable to validate these associations and to provide a definitive explanation of the underlying mechanisms. Large-scale genomic approaches, such as targeted exome and whole genome sequencing (WGS), offer promising avenues for uncovering novel candidate genes and clarifying the biological mechanisms of host susceptibility and host-pathogen interactions. These insights may ultimately pave the way for targeted interventions, even though current clinical guidelines do not yet incorporate these genomic findings [32], [33]. With increased power in future studies, GWAS or WGS approaches could be employed for host-pathogen interaction analyses that reveal avenues for targeted interventions.

Through genomic analysis as part of a national multicenter cohort study for culture-proven bacterial sepsis in children in Switzerland, we identified a genomic region significantly associated with individual susceptibility to sepsis. This offers a potential route for improving the prognosis for those affected and uncovering pathogenic mechanisms involved in immune responses and host-pathogen interactions.

## Conclusion

This genome-wide analysis in the Swiss Pediatric Sepsis Study identified a significantly associated locus on chromosome 9, centred around *CTNNAL1* and *ELP1*, considered to modulate paediatric sepsis susceptibility. These findings provide a potential avenue for improving prognosis and shed light on the underlying host-pathogen interactions, offering a foundation for future targeted interventions.

## Data Availability

All summary statistic data are present in the work. Raw genomic data is stored under restricted access according to ethics agreements. All code is published.

## Abbreviations

Actl7a and Actl7b: actin-like 7a and 7b
CC: Collaborative Cross
CTNNAL1: catenin alpha-like 1 gene
CVAD: central venous access device
DALYs: Disability Adjusted Life Years
DEG: differentially expressed gene
ELP1: elongator complex protein 1
eQTL: expression Quantitative Trait Locus
EUCLIDS: EU Childhood Life-threatening Infectious Disease Study
FAM206A: family with sequence similarity 206, member A
GRM: genetic relationship matrix
GWAS: genome-wide association study
HWE: Hardy-Weinberg Equilibrium
IBS: identify-by-state
ELP1: elongator acetyltransferase complex subunit 1
IKK: IkB enzyme kinase
LD: linkage disequilibrium
LOCO: leave-one-chromosome-out
MLMA: mixed linear model based asso-ciation
MOF: multiorgan failure
NICU: neonatal intensive care unit
PC: principal component
PCA: principal component analysis
PICU: Pediatric Intensive Care Unit
SNP: single nucleotide polymorphism
sQTL: splicing Quantitative Trait Locus
WGS: whole genome sequencing

## Weblinks

FinnGen https://r4.finngen.fi

FUMA https://fuma.ctglab.nl

GCTA https://cnsgenomics.com/software/gcta/

GTEx https://www.gtexportal.org/home/

HaploReg https://pubs.broadinstitute.org/mammals/haploreg/haploreg.php

IMPUTE2 https://mathgen.stats.ox.ac.uk/impute/impute_v2.html

KING https://people.virginia.edu/~wc9c/KING/

LDLink https://ldlink.nci.nih.gov

LocusZoom http://locuszoom.org

PLINK http://zzz.bwh.harvard.edu/plink/

SHAPEIT2 https://mathgen.stats.ox.ac.uk/genetics_software/shapeit/shapeit.html

## Contributors

Dylan Lawless performed the genomic analysis and wrote the manuscript. Flavia Aurelia Hodel, Zhi Ming Xu, Christian W. Thorball, Alessandro Borghesi, Eric Giannoni, Johannes Trück, Martin Stocker, Klara M Posfay-Barbe, Ulrich Heininger, Sara Bernhard-Stirnemann, Anita Niederer-Loher, Christian R. Kahlert, Giancarlo Natalucci, Christa Relly, Christoph Berger, Thomas Riedel, Christoph Aebi, and Philipp Agyeman contributed to data collection and manuscript drafting. Jacques Fellay and Luregn J. Schlapbach led the project and supervised the study.

## Declaration of Interest

The authors declare no conflict of interest.

## Data Sharing Statement

Summary statistic data is available from the public repository. Access to individual-level genetic data is restricted due to research use agreements. Researchers may contact the corresponding authors (L.J.S. and J.F.) to request access, subject to institutional and ethical approvals.

## Acknowledgements

We gratefully acknowledge the participation of all volunteers, and thank the medical centres and staff for their contribution.

## Supplemental

**Table S1: SNP quality control filters.**

Samples were genotyped using Illumina OmniExpressExome-8 v1.4 genotyping array, phased with (SHAPEIT2) and imputed (IMPUTE2) using the 1000 Genomes Project phase 3 reference panel. Filtering included missing genotype call rate 10%, minor allele frequencies < 0.05, control HWE 1e^−6^, and subject independence by identify-by-state (IBS). Geno, genotype call rate; HWE, Hardy-Weinberg Equilibrium; MAF, minor allele frequency; LD, linkage disequilibrium.

**Table S2: Association model summary.**

Covariates were included to account for age, sex, study sites. Ancestry was accounted for by GRM. Case-only analysis was also performed for key criteria of clinical characteristics that may be used to assess sepsis.

**Table S3. List of protein-altering variants in LD with lead SNP**

Genomic coordinates are based on the GRCh 37 build. LD estimates are based on the HRC reference panel. Haploreg v4 was used to assess the protein-altering functional annotation of the proxy SNPs.

**Table S4. Extended list of all variants in LD with lead SNP**

The extended list of variants in LD with the lead SNP contain 1519 variants, with ranges in their MAF 0.1-0.5; D’ 0.1-1.0; and R^2^ 0.0-1.0.

**Table S5. eQTL associations for associated SNPs in GTEx v8**

Genome wide significant SNPs in sepsis susceptibility case-control analysis were used to query for eQTL associations in GTEx v8. Results for the lead SNP rs28361152 are shown. P-values included are below the gene-level significant threshold of variant-gene pairs. Genomic coordinates are based on the GRCh 38 build. NES, normalized effect size; sQTL, splicing Quantitative Trait Loci.

**Table S6. sQTL associations for associated SNPs in GTEx v8**

Genome wide significant SNPs in sepsis susceptibility case-control analysis were used to query for eQTL associations in GTEx v8. Results for the lead SNP rs28361152 are shown. P-values included are below the gene-level significant threshold of variant-gene pairs. Genomic coordinates are based on the GRCh 38 build. eQTL, expression Quantitative Trait Loci; NES, normalized effect size.

**Figure S1:**
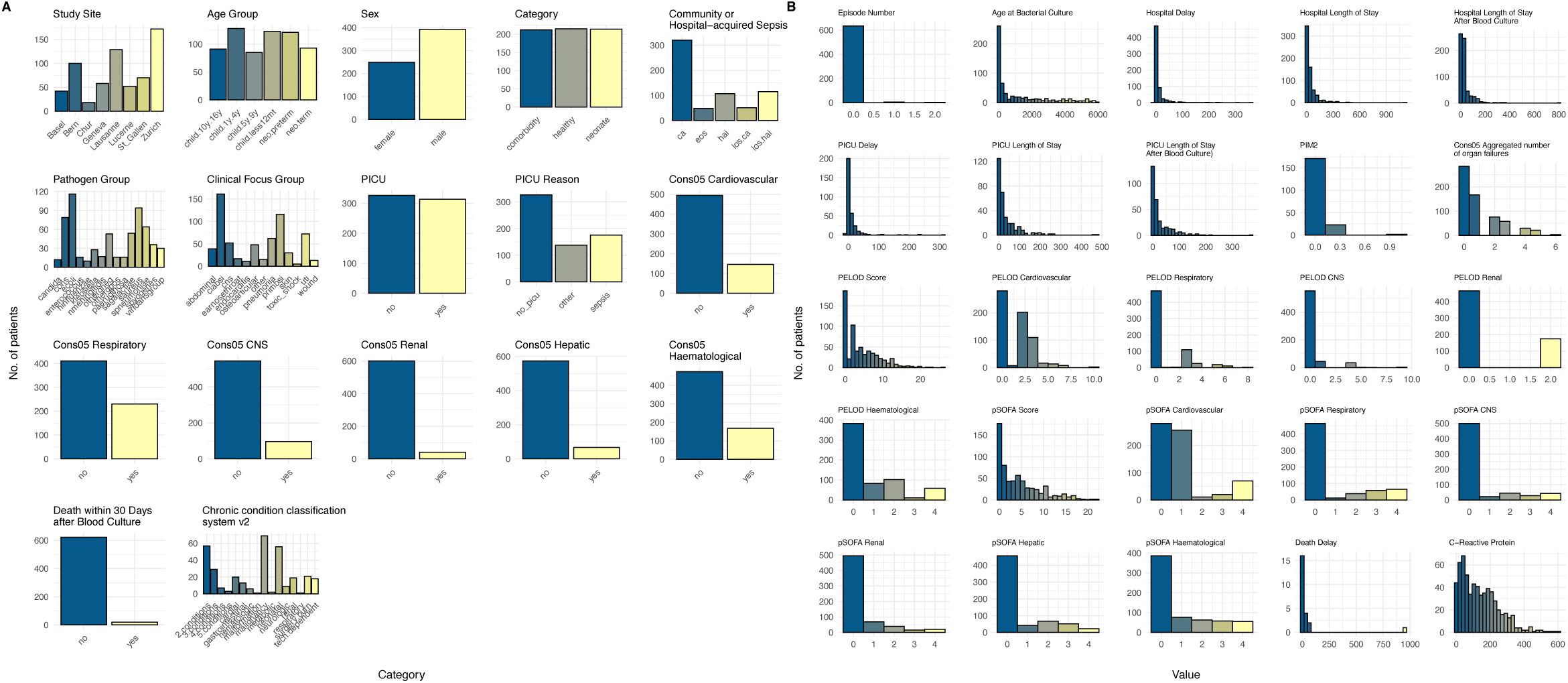
Patient cohort characteristics. (A) categorical and (B) continuous variables for patient cohort. Our cohort consisted of 650 cases after QC (quality control). Color scale low (blue) to high (yellow).

**Figure S2:**
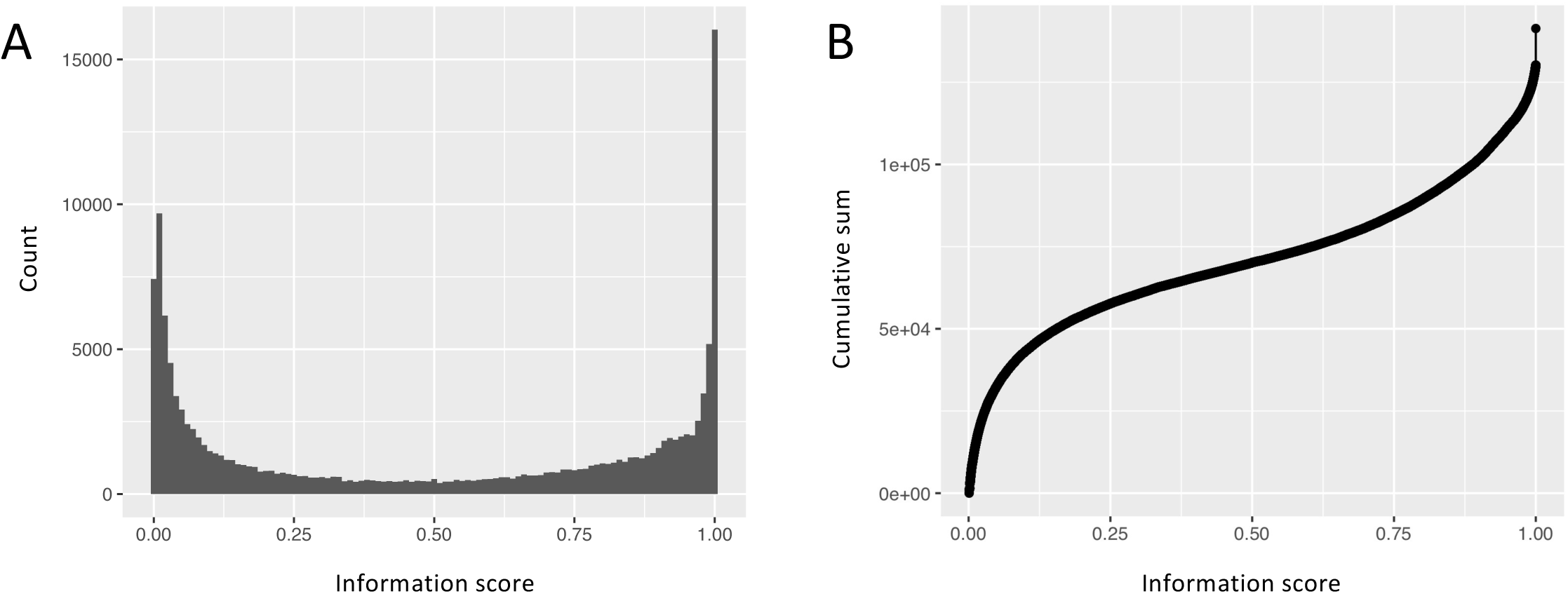
Imputation quality. The information score is plotted for illustration of a randomly selected region of 5e^6^ SNPs. High quality imputation is present for variants with scores above 0.7-0.8 as shown for (A) total count and (B) cumulative sum. INFO score >0.8 was used for analysis.

**Figure S3:**
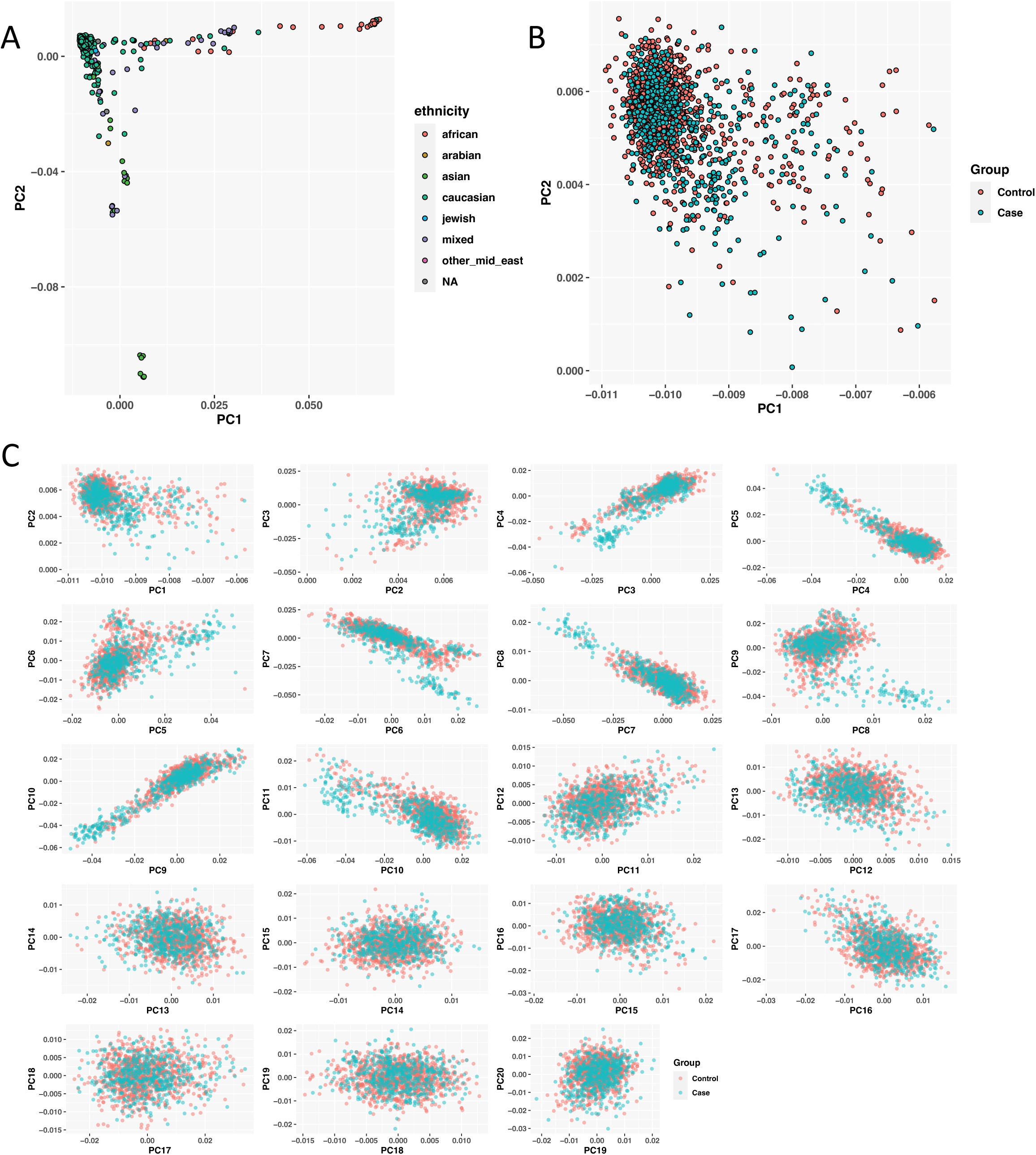
Principal component analysis. (A) The Swiss Pediatric Sepsis Study cohort is predominantly made up of individuals of European ancestry (83%, **Table 1**). Within-cohort association was tested for all samples with eight eigenvalues included as quantitative covariates to control for population structure in a mixed model. (B) For case-control analysis population structure required pruning to European only ancestry. The first twenty PCs are shown in (C).

**Figure S4:**
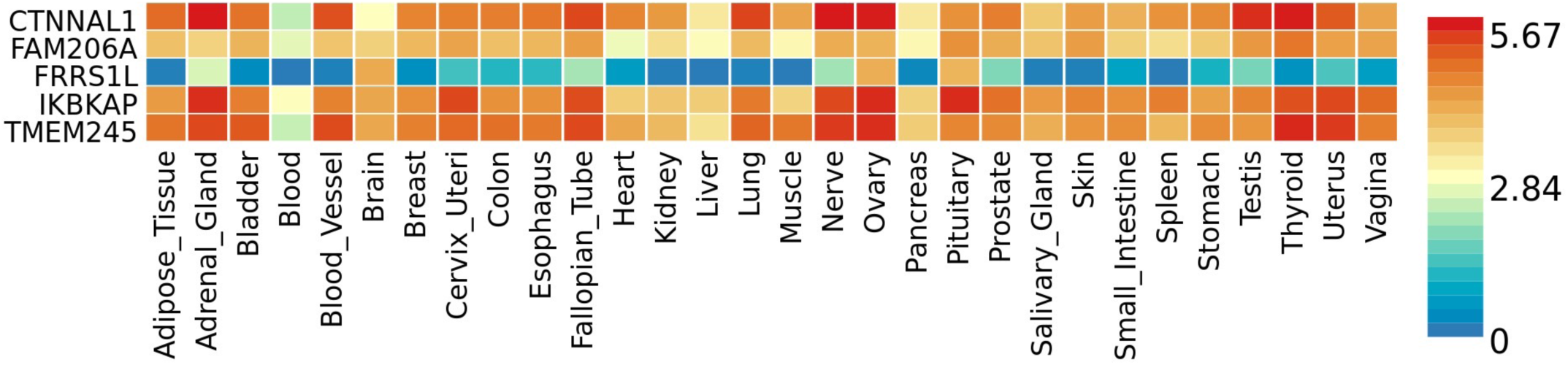
Gene expression in candidate gene locus. Gene expression heatmap from 5 genes of interest on Chr 9 in association with sepsis. Annotated for biological context based on Functional Mapping and Annotation of Genome-Wide Association Studies (FUMA). Significantly associated SNPs overlap *ELP1, FAM206A*, and *CTNNAL1* and all five genes were identified as expression Quantitative Trait Loci.

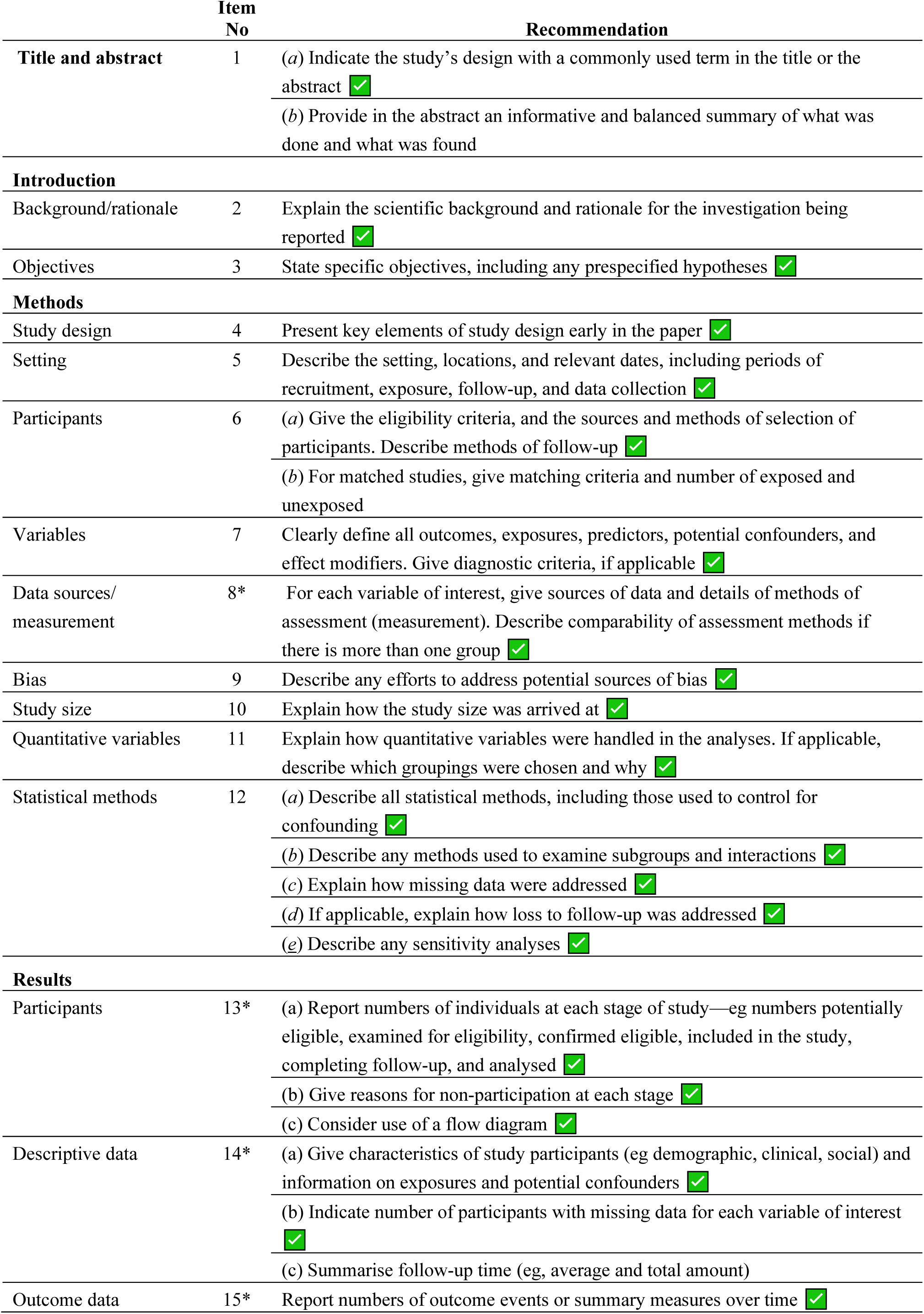
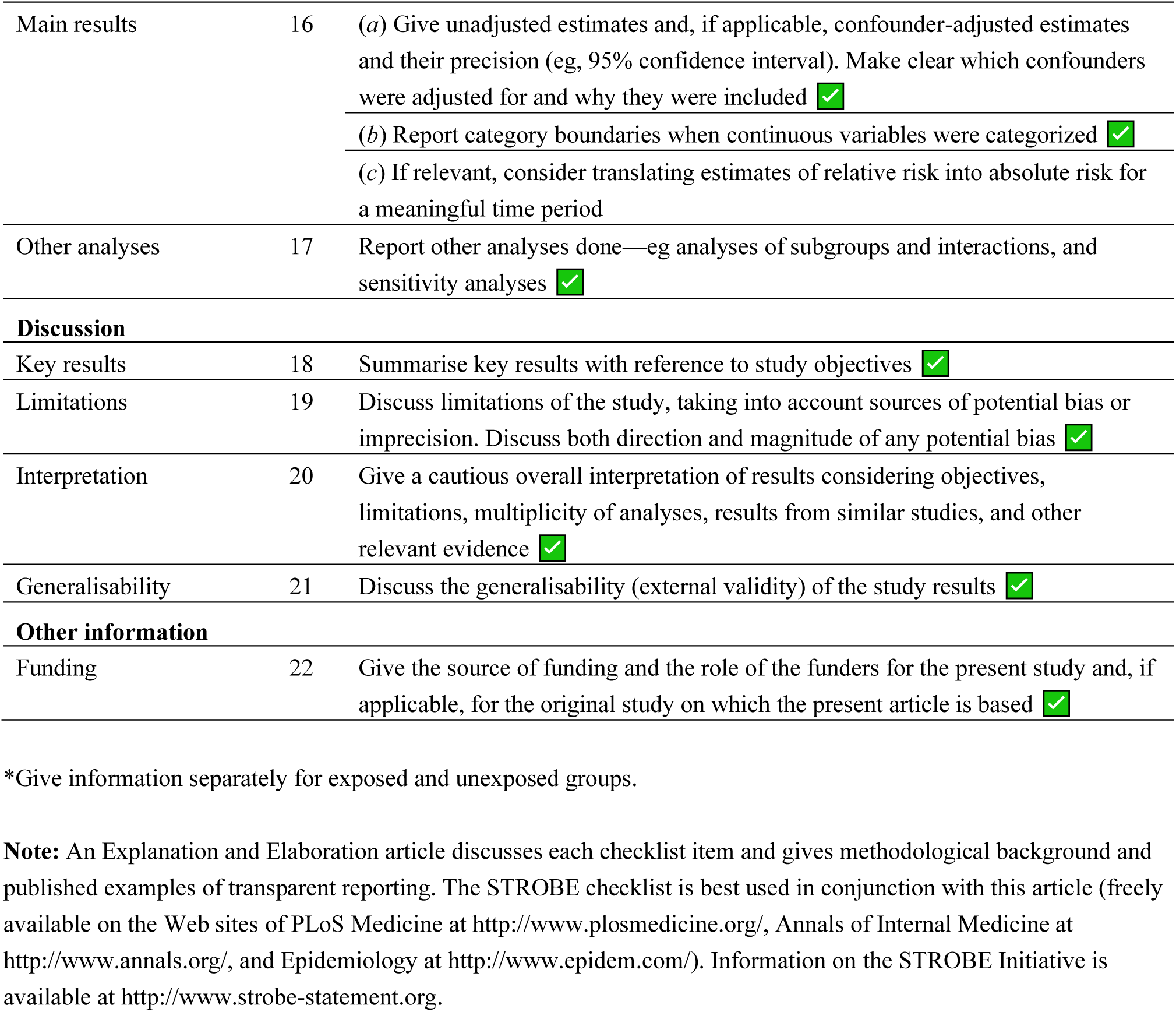
STROBE Statement—Checklist of items that should be included in reports of ***cohort studies***

|  | Item No | Recommendation |
| --- | --- | --- |
| **Title and abstract** | 1 | (a) Indicate the study's design with a commonly used term in the title or the abstract<br>(b) Provide in the abstract an informative and balanced summary of what was done and what was found |
| **Introduction** |  |  |
| Background/rationale | 2 | Explain the scientific background and rationale for the investigation being reported |
| Objectives | 3 | State specific objectives, including any prespecified hypotheses |
| **Methods** |  |  |
| Study design | 4 | Present key elements of study design early in the paper |
| Setting | 5 | Describe the setting, locations, and relevant dates, including periods of recruitment, exposure, follow-up, and data collection |
| Participants | 6 | (a) Give the eligibility criteria, and the sources and methods of selection of participants. Describe methods of follow-up<br>(b) For matched studies, give matching criteria and number of exposed and unexposed |
| Variables | 7 | Clearly define all outcomes, exposures, predictors, potential confounders, and effect modifiers. Give diagnostic criteria, if applicable |
| Data sources/<br>measurement | 8* | For each variable of interest, give sources of data and details of methods of assessment (measurement). Describe comparability of assessment methods if there is more than one group |
| Bias | 9 | Describe any efforts to address potential sources of bias |
| Study size | 10 | Explain how the study size was arrived at |
| Quantitative variables | 11 | Explain how quantitative variables were handled in the analyses. If applicable, describe which groupings were chosen and why |
| Statistical methods | 12 | (a) Describe all statistical methods, including those used to control for confounding<br>(b) Describe any methods used to examine subgroups and interactions<br>(c) Explain how missing data were addressed<br>(d) If applicable, explain how loss to follow-up was addressed<br>(e) Describe any sensitivity analyses |
| **Results** |  |  |
| Participants | 13* | (a) Report numbers of individuals at each stage of study—eg numbers potentially eligible, examined for eligibility, confirmed eligible, included in the study, completing follow-up, and analysed<br>(b) Give reasons for non-participation at each stage<br>(c) Consider use of a flow diagram |
| Descriptive data | 14* | (a) Give characteristics of study participants (eg demographic, clinical, social) and information on exposures and potential confounders<br>(b) Indicate number of participants with missing data for each variable of interest<br>(c) Summarise follow-up time (eg, average and total amount) |
| Outcome data | 15* | Report numbers of outcome events or summary measures over time |
| Main results | 16 | (a) Give unadjusted estimates and, if applicable, confounder-adjusted estimates and their precision (eg, 95% confidence interval). Make clear which confounders were adjusted for and why they were included<br>(b) Report category boundaries when continuous variables were categorized<br>(c) If relevant, consider translating estimates of relative risk into absolute risk for a meaningful time period |
| Other analyses | 17 | Report other analyses done—eg analyses of subgroups and interactions, and sensitivity analyses |
| **Discussion** |  |  |
| Key results | 18 | Summarise key results with reference to study objectives |
| Limitations | 19 | Discuss limitations of the study, taking into account sources of potential bias or imprecision. Discuss both direction and magnitude of any potential bias |
| Interpretation | 20 | Give a cautious overall interpretation of results considering objectives, limitations, multiplicity of analyses, results from similar studies, and other relevant evidence |
| Generalisability | 21 | Discuss the generalisability (external validity) of the study results |
| **Other information** |  |  |
| Funding | 22 | Give the source of funding and the role of the funders for the present study and, if applicable, for the original study on which the present article is based |

\*Give information separately for exposed and unexposed groups.
**Note:** An Explanation and Elaboration article discusses each checklist item and gives methodological background and published examples of transparent reporting. The STROBE checklist is best used in conjunction with this article (freely available on the Web sites of PLoS Medicine at http://www.plosmedicine.org/, Annals of Internal Medicine at http://www.annals.org/, and Epidemiology at http://www.epidem.com/). Information on the STROBE Initiative is available at http://www.strobe-statement.org.

## Notes

### Competing Interest Statement

The authors have declared no competing interest.

### Author Declarations

The study was approved by the respective ethics committees of all participating centers (Cantonal Ethics Committee Bern, approval number KEK-029/11) and the study was conducted in accordance with the Declaration of Helsinki.

### Summary of Updates

Formatted to include ethics and funder information in the methods, funding statement in the structured abstract, and a "Research in context" summary.

